# The Association of Alpha Globin Gene Copy Number with Exhaled Nitric Oxide in a Cross-sectional Study of Healthy Black Adults

**DOI:** 10.1101/2022.10.09.22280804

**Authors:** A. Parker Ruhl, Jarrett M. Jackson, Carlos J. Carhuas, Jessica G. Niño de Rivera, Michael P. Fay, J. Brice Weinberg, Loretta G. Que, Hans C. Ackerman

## Abstract

**Introduction:** The genetic determinants of fractional exhalation of nitric oxide (FeNO), a marker of lung inflammation, are understudied in Black individuals. Alpha globin (*HBA*) restricts nitric oxide signaling in arterial endothelial cells via interactions with nitric oxide synthase (NOS); however, its role in regulating the release of NO from respiratory epithelium is less well understood. We hypothesized that an *HBA* gene deletion, common among Black individuals, would be associated with higher FeNO.

**Methods:** Healthy Black adults were enrolled at four study sites in North Carolina from 2005-2008. FeNO was measured in triplicate using a nitric oxide analyzer. The −3.7 kb *HBA* gene deletion was genotyped using droplet digital PCR on genomic DNA. The association of FeNO with *HBA* copy number was evaluated using multivariable linear regression employing a linear effect of *HBA* copy number and adjusting for age, sex, and serum IgE levels. Post-hoc analysis employing a recessive mode of inheritance was performed.

**Results:** 895 individuals were in enrolled in the study and 720 consented for future genetic research; 643 had complete data and were included in this analysis. Median (25^th^, 75^th^) FeNO was 20 (13, 31) ppb. *HBA* genotypes were: 30 (4.7%) -a/-a, 197 (30.6%) -a/aa, 405 (63%) aa/aa, and 8 (1.2%) aa/aaa. Subjects were 35% male with median age 20 (19, 22) years. Multivariable linear regression analysis revealed no association between FeNO and *HBA* copy number (β = −0.005 [95% CI: −0.042, 0.033], p=0.81). In the post-hoc sensitivity analysis, homozygosity for the HBA gene deletion was associated with higher FeNO (β = 0.107 [95% CI 0.003, 0.212]; p = 0.045)

**Conclusion:** We found no association between *HBA* copy number and FeNO using a prespecified additive genetic model. However, a post hoc recessive genetic model found FeNO to be higher among subjects homozygous for the *HBA* deletion.

**KEY MESSAGES:**

- **What is already known on this topic** – The alpha subunit of hemoglobin restricts the release of nitric oxide from vascular endothelial cells, but alpha globin’s role in restricting exhaled nitric oxide is less well understood.
- **What this study adds** – We examined the association between fractional exhaled nitric oxide and deletion of the alpha globin gene, a frequent genetic polymorphism among people with African or Asian ancestry. We found that healthy Black individuals who were homozygous for the alpha globin gene deletion had higher fractional exhaled nitric oxide, consistent with alpha globin’s role as a nitric oxide restrictor.
- **How this study might affect research, practice or policy** – An alpha globin gene deletion, commonly found among Black individuals, may increase fractional exhaled nitric oxide. Whether this changes the normal range of fractional exhaled nitric oxide for Black individuals or impacts the risk of developing asthma requires further study.

## INTRODUCTION

Nitric oxide (NO) has numerous biological activities in the lung, serving as a bronchodilator, vasodilator, neurotransmitter, and inflammatory mediator. In asthmatic individuals, the fractional exhalation of nitric oxide (FeNO) is a validated measure of airway inflammation;(1,2) however, little is known about genetic factors influencing FeNO, particularly among healthy Black individuals.

Recently, a new paradigm of NO regulation in the vasculature has emerged in which endothelial alpha globin interacts with endothelial NO synthase (eNOS) to limit the release of NO within small arteries.(3) Genetic deletion of alpha globin, common among Black Americans, is associated with improved NO-mediated vascular perfusion and with protection from kidney disease.(4–6) Alpha globin, beta globin, and eNOS are expressed in airway epithelium and while beta globin has been found to interact directly with eNOS to regulate the oxidation of NO, the role of alpha globin in healthy respiratory epithelium remains undefined.(7–10) NO is produced by inducible NOS (iNOS) in airway epithelial and inflammatory cells(11). While the interaction of alpha globin with iNOS has not previously been studied in epithelial cells, the structure of iNOS is nearly identical to that of eNOS, including at the sites where eNOS interacts with alpha globin.(12) This raised the question of whether alpha globin regulates NO release from the respiratory epithelium. To address this question, we analyzed the association between FeNO and *HBA* copy number in healthy Black adults.

## METHODS

Healthy black individuals aged 18-40 years were enrolled in a multi-center, cross-sectional cohort at four university sites near Durham, North Carolina from 2005 to 2008.(13) All participants provided oral and written informed consent. The protocol was approved by the Duke University Institutional Review Board (#Pro00004947). Age, sex, race, and ethnicity were self-reported. Only non-Hispanic African American participants were enrolled. Participants were asked to confirm they were healthy (i.e., no chronic illnesses or chronic use of any medication except oral contraceptives); had no history of asthma, allergic rhinitis, hay fever, or atopic dermatitis; and were nonsmokers. Blood samples were obtained. *HBA* copy number was measured by droplet digital PCR (ddPCR) on genomic DNA. Total serum immunoglobulin-E (IgE) was measured using the Pharmacia CAP system. FeNO was measured in triplicate with a Sievers 280i Nitric Oxide Analyzer (GE Analytical Instruments, Boulder, Colo) according to American Thoracic Society recommendations.(1) A 50 mL/s flow rate was established against resistance to maintain 5 cmH_2_O oropharyngeal pressure.(13) Additional exclusion criteria for this analysis were: not consenting to future research and serum cotinine level ≥25 ng/mL signifying active tobacco use.

### Statistical methods

For continuous measures, medians and 25^th^ and 75^th^ percentiles were calculated by *HBA* genotype. Group differences were assessed by Kruskal-Wallis test. Categorical variables were calculated as percentages and differences were assessed by Fisher’s exact test. IgE and FeNO were log transformed due to skewness. The association of *HBA* genotype with FeNO was evaluated using multivariable linear regression employing a linear effect of *HBA* gene copy number with adjustment for age, sex, and total serum IgE levels. BMI was previously found not to be associated with FeNO in this cohort and was not included in the model.(13) Two post-hoc sensitivity analyses were performed: one evaluated *HBA* copy number as a categorical variable and one evaluated a recessive mode of inheritance in which the homozygous deletion genotype (-a/-a) was compared against all other genotypes (-a/aa, aa/aa, and aa/aaa).

### Patient and Public Involvement

Patients or members of the public were not included in the conceptual design of the original study completed in 2008. We plan to discuss our research findings with members of the public who may be enrolled in other studies of genetic globin variants. In addition, we recognize the importance of involving patients and the public in the conceptual design of future studies.

## RESULTS

Of 895 original study participants, 720 consented for future research and had DNA available for genotyping. Sixty-four participants were excluded due to high cotinine levels and 13 were excluded due to indeterminate *HBA* genotype. The remaining 643 participants were 35% male and had a median (25^th^,75^th^) age of 20 (19, 22) years, serum IgE level of 58.3 (22, 160) kU/L, and FeNO value of 20 (13, 31) ppb (Table 1).

**Table 1.**
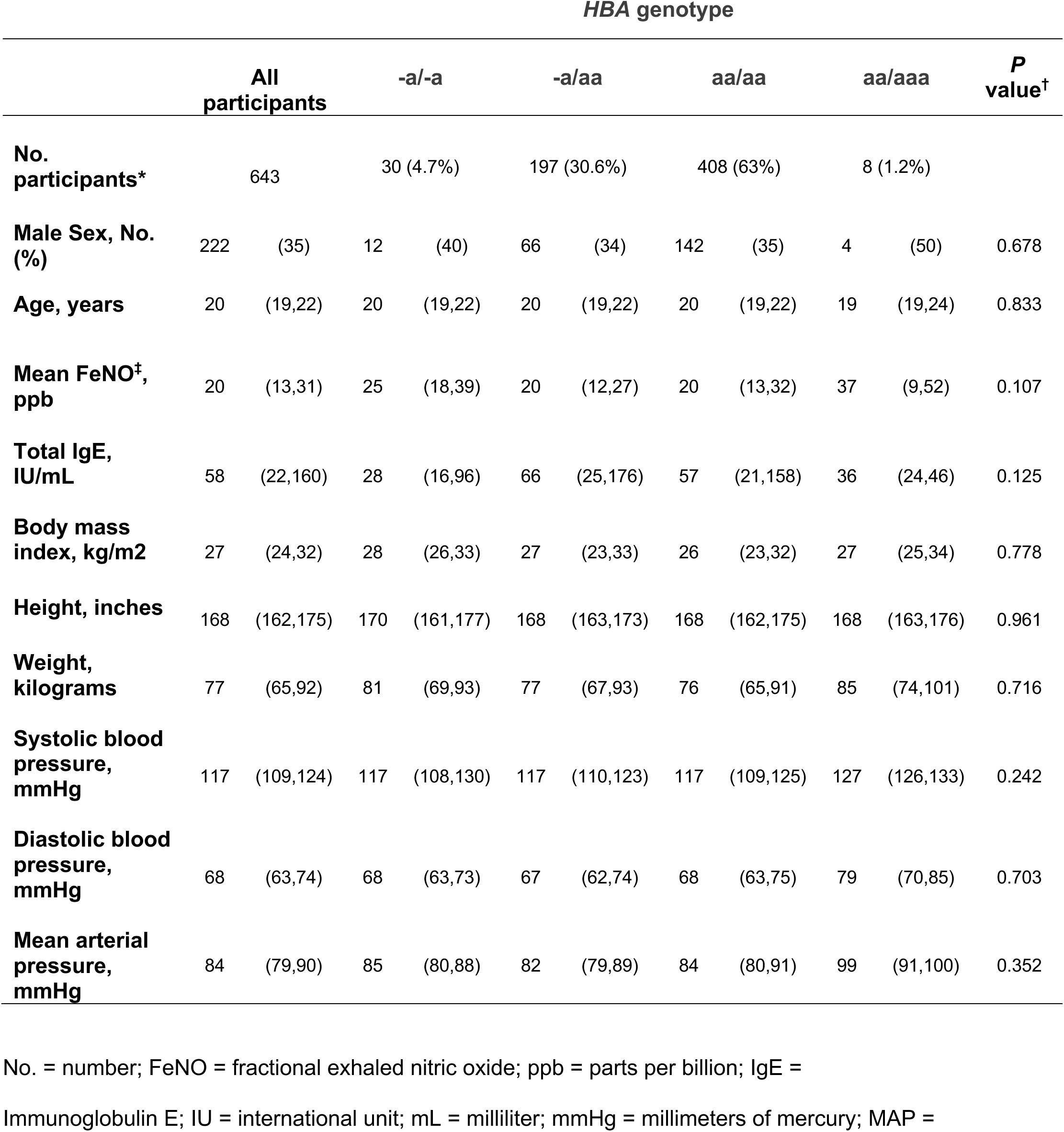
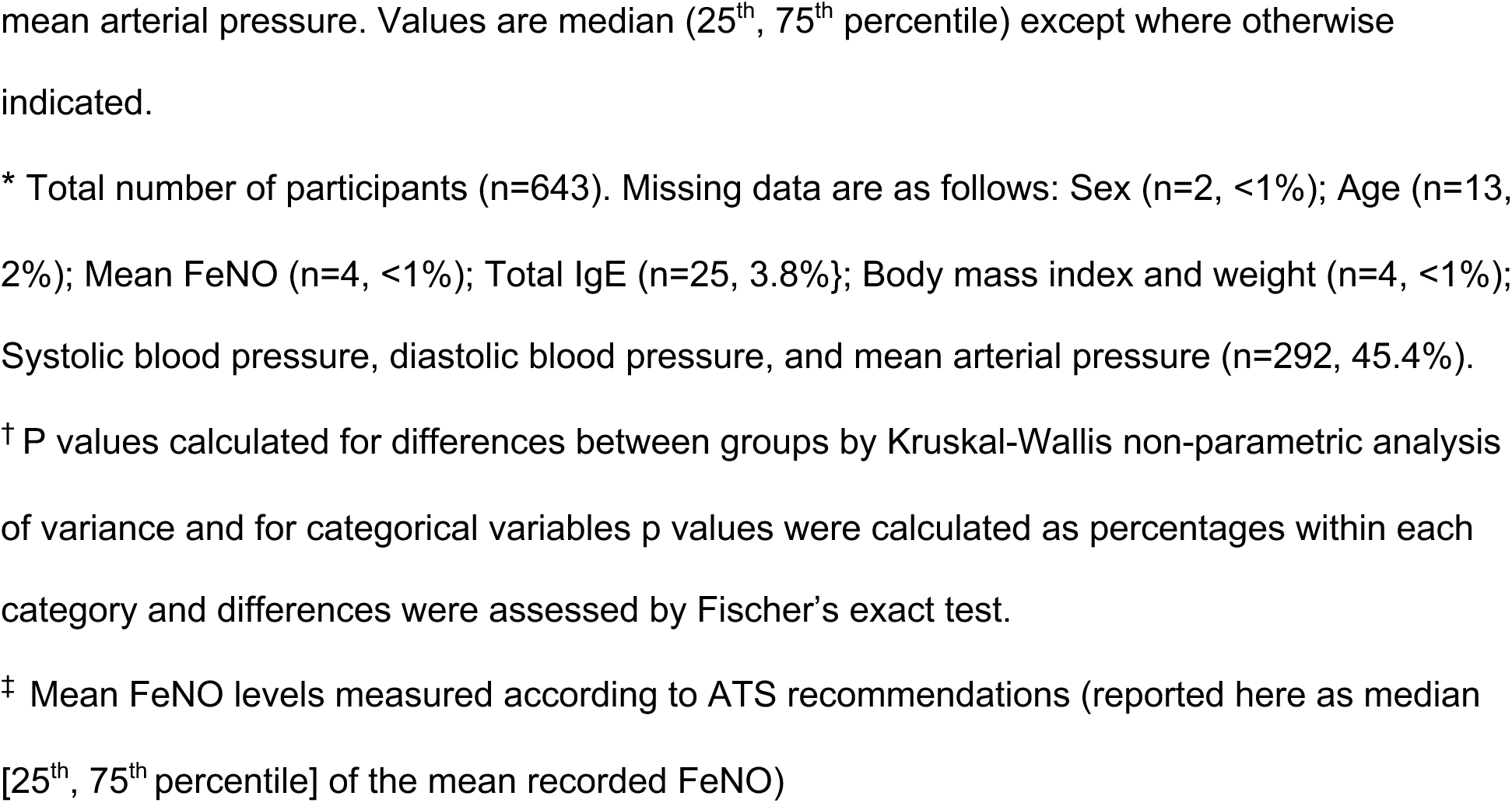
Participant characteristics grouped by alpha globin genotype.

| *HBA* genotype |  |  |  |  |  |  |  |  |  |  |  |
| --- | --- | --- | --- | --- | --- | --- | --- | --- | --- | --- | --- |
|  | All participants |  | -a/-a |  | -a/aa |  | aa/aa |  | aa/aaa |  | *P* value<sup>†</sup> |
| No. participants* | 643 |  | 30 (4.7%) |  | 197 (30.6%) |  | 408 (63%) |  | 8 (1.2%) |  |  |
| Male Sex, No. (%) | 222 | (35) | 12 | (40) | 66 | (34) | 142 | (35) | 4 | (50) | 0.678 |
| Age, years | 20 | (19,22) | 20 | (19,22) | 20 | (19,22) | 20 | (19,22) | 19 | (19,24) | 0.833 |
| Mean FeNO<sup>‡</sup>, ppb | 20 | (13,31) | 25 | (18,39) | 20 | (12,27) | 20 | (13,32) | 37 | (9,52) | 0.107 |
| Total IgE, IU/mL | 58 | (22,160) | 28 | (16,96) | 66 | (25,176) | 57 | (21,158) | 36 | (24,46) | 0.125 |
| Body mass index, kg/m2 | 27 | (24,32) | 28 | (26,33) | 27 | (23,33) | 26 | (23,32) | 27 | (25,34) | 0.778 |
| Height, inches | 168 | (162,175) | 170 | (161,177) | 168 | (163,173) | 168 | (162,175) | 168 | (163,176) | 0.961 |
| Weight, kilograms | 77 | (65,92) | 81 | (69,93) | 77 | (67,93) | 76 | (65,91) | 85 | (74,101) | 0.716 |
| Systolic blood pressure, mmHg | 117 | (109,124) | 117 | (108,130) | 117 | (110,123) | 117 | (109,125) | 127 | (126,133) | 0.242 |
| Diastolic blood pressure, mmHg | 68 | (63,74) | 68 | (63,73) | 67 | (62,74) | 68 | (63,75) | 79 | (70,85) | 0.703 |
| Mean arterial pressure, mmHg | 84 | (79,90) | 85 | (80,88) | 82 | (79,89) | 84 | (80,91) | 99 | (91,100) | 0.352 |

No. = number; FeNO = fractional exhaled nitric oxide; ppb = parts per billion; IgE =
Immunoglobulin E; IU = international unit; mL = milliliter; mmHg = millimeters of mercury; MAP =
mean arterial pressure. Values are median (25<sup>th</sup>, 75<sup>th</sup> percentile) except where otherwise indicated.
\* Total number of participants (n=643). Missing data are as follows: Sex (n=2, <1%); Age (n=13, 2%); Mean FeNO (n=4, <1%); Total IgE (n=25, 3.8%); Body mass index and weight (n=4, <1%); Systolic blood pressure, diastolic blood pressure, and mean arterial pressure (n=292, 45.4%).
† P values calculated for differences between groups by Kruskal-Wallis non-parametric analysis of variance and for categorical variables p values were calculated as percentages within each category and differences were assessed by Fischer's exact test.
‡ Mean FeNO levels measured according to ATS recommendations (reported here as median [25<sup>th</sup>, 75<sup>th</sup> percentile] of the mean recorded FeNO)

*HBA* deletion was common with 30 (4.7%) -a/-a, 197 (30.6%) -a/aa, 405 (63%) aa/aa, and 8 (1.2%) aa/aaa genotypes. Median (25^th^, 75^th^) FeNO was 25 (18, 39) ppm in the -a/-a group, 20 (12, 27) ppm in the -a/aa group, 20 (13, 32) ppm in the aa/aa group, and 37 (9, 52) ppm in the aa/aaa group (Table 1 and Figure 1). In an unadjusted linear regression analysis using the pre-specified additive genetic model, the coefficient for *HBA* copy number with FeNO was 0.001 (95% CI: −0.039, 0.040; p=0.978). After adjustment for sex, age, and serum IgE, the coefficient for *HBA* copy number with FeNO was −0.005 (95% CI: −0.042, 0.033; p=0.811; Table 2). In post hoc sensitivity analyses, the adjusted association between the homozygous genotype -a/-a and FeNO was 0.099 (95% CI −0.007, 0.206; p = 0.066) when analyzed as a categorical variable and 0.107 (95% CI 0.003, 0.212; p = 0.045; Table 2) when analyzed using a recessive mode of inheritance.

**Figure 1.**
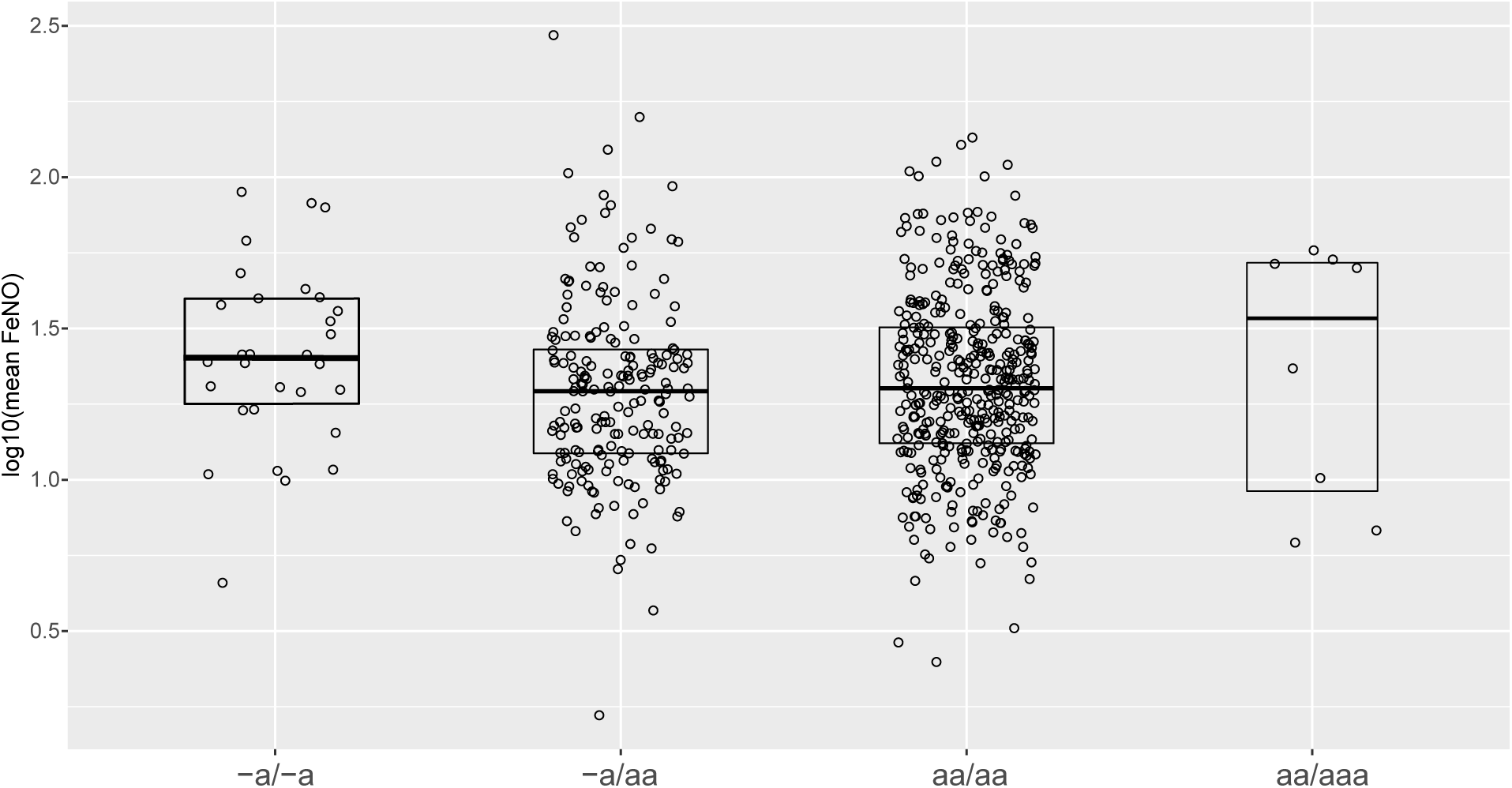
Fractional exhaled nitric oxide levels in 643 Black individuals grouped by alpha globin genotype.

**Table 2.** Multivariable regression analysis of *HBA* genotype and fractional exhaled nitric oxide*.

| **Multivariable linear regression model employing *HBA* genotype as an integer gene copy number** |  |  |  |
| --- | --- | --- | --- |
|  | Beta Coefficient | 95% Confidence Interval | *P* value |
| *HBA* copy number, per copy | -0.005 | (-0.042, 0.033) | 0.811 |
| Age, per year | -0.001 | (-0.006, 0.003) | 0.587 |
| Male sex | 0.122 | (0.074, 0.170) | < 0.001 |
| Log<sub>10</sub> IgE | 0.137 | (0.098, 0.176) | < 0.001 |
| **Post hoc multivariable linear regression model employing *HBA* genotype with a recessive mode of inheritance** |  |  |  |
|  | Beta Coefficient | 95% Confidence Interval | *P* value |
| *HBA* genotype, -a/-a | 0.107 | (0.003, 0.212) | 0.045 |
| Age, per year | -0.001 | (-0.006, 0.003) | 0.541 |
| Male sex | 0.120 | (0.071, 0.166) | < 0.001 |
| Log<sub>10</sub> IgE | 0.140 | (0.102, 0.181) | <0.001 |

*HBA* = alpha globin gene; No. = number; Log<sub>10</sub> = log base 10; IgE = immunoglobulin E
\* Out of n=643 participants contributing data to the multivariable regression models n=44 participants were excluded due to missing data and n=599 participants were included in the multivariable regression models. Missing data by variable are as follows: Age (n=13, 2%); Sex (n=2, <1%); Total IgE (n=25, 3.8%); Mean FeNO (n=4, <1%).

## DISCUSSION

Alpha and beta globin have recently emerged as regulators of NO signaling in the vascular endothelium and respiratory epithelium, respectively;(3,10) however, there are few studies evaluating the impact of globin gene variants on NO signaling in vivo. In this study, we characterized a common *HBA* deletion in healthy Black individuals and examined the relationship between *HBA* copy number and FeNO. We found no association between *HBA* genotype and FeNO using a pre-specified additive genetic model; however, a post hoc analysis using a recessive mode of inheritance identified homozygosity for the *HBA* gene deletion to be associated with higher FeNO levels. This latter finding is consistent with the proposed mechanism that alpha globin limits the release of nitric oxide and suggests that lower alpha globin expression allows greater release of NO from respiratory epithelium in healthy, non-asthmatic individuals. More work is needed to understand the role of epithelial alpha globin in the setting of inflammatory lung disease and to determine whether alpha globin interacts with iNOS, which is structurally similar to eNOS, and is expressed under allergic or inflammatory conditions.(12,14)

Study strengths included the large cohort size, representation of an understudied minority population, high frequency of the *HBA* gene deletion, and a well-defined quantitative outcome measure. Adjustment for serum IgE, which is associated with FeNO, was a strength of this study; however, the absence of evaluation for subclinical IgE sensitization was a limitation. Other limitations included post hoc testing of different genetic inheritance models, performing FeNO measurement at a single flow rate that does not distinguish alveolar from bronchial NO sources,(15) and absence of data on other factors that influence FeNO such as recent upper respiratory tract infection, eosinophilic cationic protein, eosinophil count, neutrophil count, and other inflammatory markers such as periostin.

The genetic epidemiological approach we used relies on variation in the alpha globin gene, which has previously been associated with vascular NO signaling and vascular disease risk, but has not previously been studied in relation to exhaled NO. Our approach did not distinguish between iNOS or eNOS as the source of NO. Our observation of a significant association with alpha thalassemia trait (two alpha globin deletions) and increased FeNO, may be consistent with prior epidemiological studies that noted a positive association with thalassemia and asthma in Taiwanese children.(16) Future studies are needed to better understand the mechanisms by which globins regulate NO signaling in the human respiratory epithelium and whether genetic variation in epithelial globin expression influences asthma risk across different ethnic groups, including those with African ancestry and southeast Asian ancestry.

## CONCLUSIONS

*HBA* copy number was not associated with FeNO among healthy Black adults when analyzed using a prespecified additive genetic model. However, FeNO was found to be higher among subjects who were homozygous for the *HBA* deletion in a post hoc analysis employing a recessive mode of inheritance. This relationship between *HBA* copy number and exhaled nitric oxide and merits further study in other populations where alpha globin is polymorphic. The mechanistic link between alpha globin and nitric oxide synthase warrants further characterization to better understand the impact of human genetic variation in alpha globin on respiratory function and health.

## Data Availability

Deidentified data and statistical code from this article will be available to researchers. Data can be obtained upon reasonable request by contacting the corresponding author.

## ACKNOWLEDGEMENTS

Several co-authors have new current affiliations, Jarrett Jackson is at the Vanderbilt University Medical Center, Nashville, TN, United States; Carlos Carhuas is at the Comprehensive Sickle Cell Disease Program, Children’s National Medicine Center, Washington, DC, United States; and Jessica Nino De Rivera is at the Columbia University Vagelos College of Physicians and Surgeons, New York, NY, United States

## Author contributions

All listed authors meet ICJME recommendations for authorship criteria, including substantial contributions to the conception or design of the work (A.P.R., H.C.A., L.G.Q., J.B.W.); or the acquisition, analysis, or interpretation of data for the work (A.P.R., H.C.A., J.M.J., C.J.C., J.G.N., M. P. F., L.G.Q.); Drafting the work (A.P.R.) or revising it critically for important intellectual content and final approval of the version to be published (All).

## REFERENCES

1. Khatri SB, Iaccarino JM, Barochia A, Soghier I, Akuthota P, Brady A, et al. Use of Fractional Exhaled Nitric Oxide to Guide the Treatment of Asthma: An Official American Thoracic Society Clinical Practice Guideline. Am J Respir Crit Care Med. 2021 Nov 15;204(10):e97– 109.

2. Jeppegaard M, Veidal S, Sverrild A, Backer V, Porsbjerg C. Validation of ATS clinical practice guideline cut-points for FeNO in asthma. Respir Med. 2018 Nov;144:22–9.

3. Straub AC, Lohman AW, Billaud M, Johnstone SR, Dwyer ST, Lee MY, et al. Endothelial cell expression of haemoglobin α regulates nitric oxide signalling. Nature. 2012 Nov 15;491(7424):473–7.

4. Denton CC, Shah P, Suriany S, Liu H, Thuptimdang W, Sunwoo J, et al. Loss of alpha-globin genes in human subjects is associated with improved nitric oxide-mediated vascular perfusion. Am J Hematol. 2020 Nov 28;

5. Romana M, Reminy K, Moeckesch B, Charlot K, Hardy-Dessources MD, Doumdo L, et al. Loss of alpha globin genes is associated with improved microvascular function in patients with sickle cell anemia. American Journal of Hematology. 2021;96(5):E165–8.

6. Ruhl AP, Jeffries N, Yang Y, Naik RP, Patki A, Pecker LH, et al. Alpha Globin Gene Copy Number Is Associated with Prevalent Chronic Kidney Disease and Incident End-Stage Kidney Disease among Black Americans. JASN. 2022 Jan 1;33(1):213–24.

7. Bhaskaran M, Chen H, Chen Z, Liu L. Hemoglobin is expressed in alveolar epithelial type II cells. Biochemical and Biophysical Research Communications. 2005 Aug 12;333(4):1348– 52.

8. Newton DA, Rao KMK, Dluhy RA, Baatz JE. Hemoglobin Is Expressed by Alveolar Epithelial Cells. J Biol Chem. 2006 Mar 3;281(9):5668–76.

9. Grek CL, Newton DA, Spyropoulos DD, Baatz JE. Hypoxia Up-Regulates Expression of Hemoglobin in Alveolar Epithelial Cells. Am J Respir Cell Mol Biol. 2011 Apr;44(4):439–47.

10. Marozkina N, Smith L, Zhao Y, Zein J, Chmiel JF, Kim J, et al. Somatic cell hemoglobin modulates nitrogen oxide metabolism in the human airway epithelium. Sci Rep. 2021 Jul 29;11(1):15498.

11. Ricciardolo FLM, Sterk PJ, Gaston B, Folkerts G. Nitric Oxide in Health and Disease of the Respiratory System. Physiological Reviews. 2004 Jul;84(3):731–65.

12. Fischmann TO, Hruza A, Niu XD, Fossetta JD, Lunn CA, Dolphin E, et al. Structural characterization of nitric oxide synthase isoforms reveals striking active-site conservation. Nat Struct Biol. 1999 Mar;6(3):233–42.

13. Levesque MC, Hauswirth DW, Mervin-Blake S, Fernandez CA, Patch KB, Alexander KM, et al. Determinants of exhaled nitric oxide levels in healthy, nonsmoking African American adults. Journal of Allergy and Clinical Immunology. 2008 Feb 1;121(2):396–402.e3.

14. Roos AB, Mori M, Grönneberg R, Österlund C, Claesson HE, Wahlström J, et al. Elevated exhaled nitric oxide in allergen-provoked asthma is associated with airway epithelial iNOS. PLoS One. 2014;9(2):e90018.

15. Linn WS, Rappaport EB, Eckel SP, Berhane KT, Zhang Y, Salam MT, et al. Multiple-flow exhaled nitric oxide, allergy, and asthma in a population of older children. Pediatric Pulmonology. 2013;48(9):885–96.

16. Piel FB, Weatherall DJ. The α-Thalassemias. New England Journal of Medicine. 2014 Nov 13;371(20):1908–16.

